# A Complex Model of Clinical Narrative Information for the Diagnostic Act

**DOI:** 10.1101/2021.02.21.21252158

**Authors:** David Chartash, Marc B Rosenman, Johan Bollen, Markus Dickinson, Stephen M Downs

## Abstract

**Background:** The act of diagnosis is one which precipitates semiotic closure, the complex integration of signs and symptoms through cognitive perspectives to ultimately activate causal reasoning and calibrate the assignment of a disease entity to the patient. In writing about this act, physicians encode both structured and unstructured information into the medical record. Unstructured information contains a latent structure which entwines both the cognitive components of the diagnostic act and the linguistic patterns associated with clinical documentation. Existing models of clinical language primarily use a physical or dialogic model of information as their basis, and do not adequately account for the complexity inherent in the diagnostic act.

**Methods:** Framing the diagnostic information collected in clinical care as a narrative, we developed a model representative of said information, accounting for its content and structure, as well as the inherent complexity therein. Using an exemplar text, we present the use of known predication and semantic relations from ontological (the Unified Medical Language System) and linguistic theory (Rhetorical Structure Theory) to facilitate the operationalization of the model, and analyze the result.

**Results:** The resulting model is demonstrated to be complex, representative of the clinical narrative text, and is fundamentally aligned with the clinical acts of both documentation and diagnosis. We find the model’s representation of the cognitive aspects of narrative consistent with models of reading, as well as an adequate model of information as presented by clinical medicine and the clinical sub-language.

**Conclusions:** We present a model to represent diagnostic information in the physician’s note which accounts for the clinical and textual narrative precipitated by the cognition involved in encoding said information into the unstructured medical record. This model prepends the development of (computational) linguistic models of the clinical sublanguage within the physician’s note as it relates to diagnosis, beyond the information level of the lexical unit. Such analysis would facilitate better reflection on the structure and meaning of the clinical note, offering improvements to medical education and care.

## Introduction

Diagnosis is an act of causal reasoning, supported by the semantic information collected during the clinical encounter. This is a definition drawn from the scientific endeavour which has become a part of modern medical practice: the systematization of diagnoses and disease. The earliest mention of this systematization is that by *Sir Thomas Clifford Allbutt* in his seminal 1896 text *“A System of Medicine”:*

> “Clinical diagnosis, however, is not investigation - a distinction some practitioners forget; diagnosis depends not upon all facts, but upon crucial facts. Indeed we may go farther and say that accumulation of facts is not science. Rather, science is our conception of the facts: the act of judgment, perhaps of imagination, by which we connect the unknown with the known.” [1]

The accumulation of facts, or the codification of a symbol space in the case of information, is not science, but rather science is the conception and judgement connecting these facts to known observation. This act of connection, or the assignment of meaning by relating symbols to reference objects, constitutes semiotic closure (sec Figure 1). In clinical parlance, this is the activation of causal logic connecting signs and symptoms to disease objects through the act of diagnosis. [2] We can therefore infer that *Allbutt* understood that the act of clinical reasoning for diagnostic purpose builds upon not only our knowledge of known diseases but also the semantic structure of clinical information and cognition.

**Figure 1.**
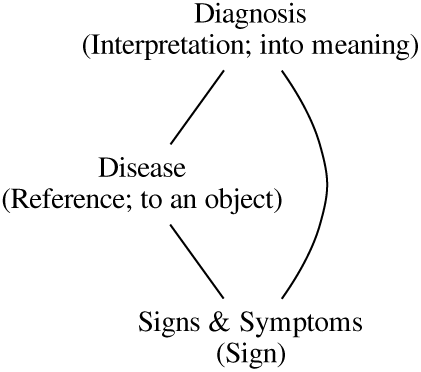
Clinical Semiotic Triad. Coupled with the standard semiotic triad in parentheticals.

Furthering this notion of semiotic closure is the act of clinical reasoning framed through cognitive perspectives. *Lemieux and Bordage* describe clinical reasoning using the cognitive science model of the mind [3]:

> “The clinician who recognizes the underlying structure of a clinical problem makes, from a propositional perspective, an appropriate selection of relevant clinical findings and, from a structural semantic perspective, an abstraction of those findings into pertinent networks of formal qualities. These two structural components interact to activate causal logic and the generation of appropriate diagnoses.” [4]

These two cognitive perspectives integrate to perform the diagnostic act, and in turn provide semiotic closure.

This duology of a propositional [5] and structural semantic perspective [6], analogizes to the two systems common to the universal model of diagnostic reasoning [7] which make up the model used in the description of clinical cognition. [8] This model of clinical cognition is the integration of two systems of reasoning: system I and system II. System I is characterized as heuristic and intuitive, associated with experimental or inductive reasoning. This reasoning parallels the structural semantic perspective in which identified findings are connected given formal qualities, given an unknown existing structure. System II is characterized as systematic and analytical, associated with hypothetico-deductive reasoning. This reasoning parallels the propositional perspective, in which relevant clinical findings are identified, given a known structure. Framing the integration of these two perspectives as the act of diagnosis, the clinical optimization found in the integration of information [9] is consistent with the model of clinical cognition’s calibration stage. The calibration stage receives inputs from both system I and II processes, such that it leads directly to diagnosis. Therefore, to model this calibration stage and the integration of the two systems, or of the two cognitive perspectives (propositional and structural semantic), is to model the integration of information and the activation of causal logic to perform the act of diagnosis. Ultimately, clinical reasoning serves as a mechanism to integrate information and activate causal logic by calibration and deliberative practice. This reasoning serves as the mechanism within the diagnostic act, and as such is part of the larger feedback system in which physicians examine the patient. [10]

This paper seeks to model the act of diagnosis, addressing the semantic and semiotic information structure of the act, rather than the Bayesian information structure pertaining to the endpoint of diagnostic classification. Specifically, this means examining the act of diagnosis through the clinical narrative from the perspective of the cognitive and linguistic framing of clinical information, rather than the socially mediated context of the clinical encounter. This is counter to the information-based efforts to model knowledge representation [11, 12] which rely on the entrenched Bayesian model of clinical reasoning [13-16] and has as an endpoint the classification task at the core of diagnostic artificial intelligence. [17-22]

## Methods

Modeling the act of diagnosis, we propose a meta-model of information sources and cognitive perspectives (diagrammed in Figure 2). Drawing on the work of *Kay and Purves* [23], this meta-model can be described as a clinical narrative given the context of the medical record. Such a clinical narrative is the combination of content and expression within the medical record, or the “what” and the “how”. While in other framings this relates to explicit references in text, in the context of the medical record this includes all information regarding a patient for an encounter (such that an encounter is the unit of time under investigation). The text of the clinical note provides a lens with which to examine the entirety of content, and goes beyond a simple summary of ancillary investigations (such as laboratory or imaging) integrated by the physician to make a diagnosis or justify a procedure. Describing the meta-model of information within the medical record as a narrative also affords for the use of narrative theory to better model the content and expression of the clinical encounter.

**Figure 2.**
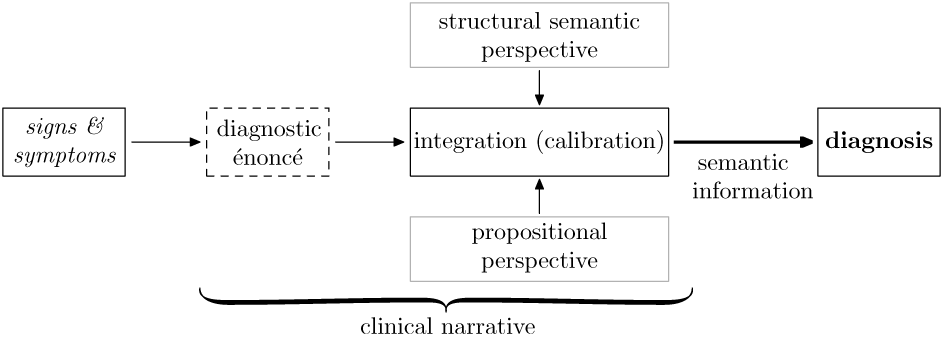
The Meta-Model of Diagnostic Information. Signs & symptoms are encoded in clinical text as diagnostic énoncés, and when integrated with the propositional and structural semantic perspectives they articulate the semantic model information leading to diagnosis. This integration is representative of the meta-model of clinical narrative. The blocks corresponding to fabula elements are indicated by a dashed black border, while those corresponding to the sjuzhet are indicated with a solid gray border.

Narrative theory [24] describes content as mostly analogous to the *fabula* and expression to the *sjuzhet*. The fabula is defined as as “the set of events and existents which make up the core of the narrative” and the sjuzhet as “the arrangement of the events into and order or structure via signification and semantics”. The description and definition of the elements of the narrative confirms that the clinical narrative models the integration of the content within the medical record through the identification of core events and their organization and structure. This integration is the intended interaction between the propositional and structural semantic perspectives, where clinical findings are selected (core events identified) and abstracted into pertinent networks (i.e. structures) to activate causal logic.

### The Diagnostic Énoncé

While philosophically we have described a clinical narrative modeling the information within the clinical record (representative of diagnostic reasoning), the nature of that information is still vague as it pertains to textual data. *Leguil* [25] describes the encoding of clinical signs and symptoms to match syndromes or disease as the act of diagnosis. These clinical signs and symptoms are polysemous, and map to more than one disease, forming an *N*-ary to *M*-ary network. This *N-*ary to *M*-ary mapping is the same mapping described by *Blois* [26] in his articulation that diagnosis is both recognition and classification as it pertains to an information theoretic representation of signs and symptoms. Furthermore, this notion of diagnostic information mapping fits with the notion that diagnosis is not solely a labeling task, but rather as an input to the clinical reasoning process. [27] The reconciliation of the *N*-ary to *M*-ary mapping is through the assignment of meaning: semiotic closure. To codify such semiotic closure, whether resultant from mapping or clinical reasoning as a process, *Leguil* describes the diagnostic énoncé. The diagnostic énoncé is the unit of medical discourse which renders the causal schematic representation of *Lantéri-Laura* [28]; polysemy connecting sign and symptom are rendered through the concept of information or index, a clinimetric in terms of *Feinstein*. [29] We can conclude, therefore, with further assistance from *Mounin* in his clarification of medical semiology [30], that the discrete entity of the clinimetric index as measured from the patient creates meaning from both internal and external relations. This duology is analogous to the propositional and structural semantic perspectives of *Lemieux and Bordage*, and provides us with a semiotic basis for using the diagnostic énoncé as the unit of discourse when modeling the two perspectives. The diagnostic énoncé provides a bridge between the medical and linguistic discourse. The énoncé, a linguistic unit of discourse, encodes the information within a textual narrative as the core element of the narrative. When facilitative of diagnostic information (and so characterized) it encodes information as the core element of the clinical narrative.

Therefore, linguistic methods can be used to describe the information within the medical record. Linguistic structures have analogues to the énoncé. *Greimas* [31, 32] describes the interaction between the elementary unit of narrative and units of language. The énoncé contains two forms of syntagm configurations (Figure 3).

**Figure 3.**
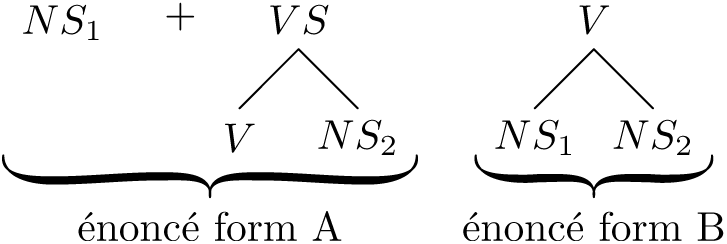
Elementary Units of Narrative. Organized into syntagam forms (énoncés) described by *Greimas*. [32]

A syntagm is defined as:

> «Groupe d’unités linguistiques significatives formant une unité dans une organisation hiérarchisée de la phrase.» [a group of significant linguistic units which form a unit within the hierarchical organization of the phrase] ^[1]^

Syntagms can therefore be taken as linguistic units which are chained together within the hierarchical organization of the phrase. In the case of Figure 3, there are two types of syntagms used within narrative: nominal and verbal. The former qualifies the syntagm as a unit intermediate between the noun-phrase and the noun at the head of the noun-phrase. The latter qualifies the syntagm as having a verb as the main element of the unit intermediate between the verb-phrase and the verb at the head of the verb-phrase. [33] *Greimas* includes the nominal syntagm as a constituent component of the sentence, and replaces it with the noun phrase. This replacement is such that the narrative énoncé can be said to be equivalent to the linguistic structure of the elementary discourse unit.

### Complexity

While the diagnostic énoncé serves as the descriptive mechanism for narrative events, the structure of the narrative is facilitated by the semantic structure present in the story. Applying the discourse theory of Rhetorical Structure Theory (RST), we can examine such structure. [34] Within the context of a span of text, RST provides a means of organization connecting elementary discourse units, énoncés, according to a set of semantic strictures (schema) and relations. For a constitutive element within the narrative, this provides a (hierarchical) system of relations between the core concept expressed by the element and the surrounding text, making the narrative non-linear as its sjuzhet connects to its fabula, as well as how each element contains a connected system of elementary discourse units.

From comparative studies of folkloric narratives, we can confirm that non-linearity is not contingent on the complexity of individual fabula elements. For example, in the story of the Princess and the Pea (ATU 704), when comparing between the German and Danish traditions, sjuzhet non-linearity is observed between the number and type of tests, as well as the agency of the prince in seeking the princess. [35] Additionally, from cognitive models of narrative, we know that the read perspective of narrative sequence is often non-linear. In constructing a model of reading, *Trabasso* [36] has suggested that a causal framework representative of a coherent sequence of events describes the read story. This framework is representative of causal, temporal and logical inferences which construct a discourse model of narrative.

Given the non-linear model of cognition inherent to medical thought and the use of knowledge in clinical reasoning [3, 37], a network model is useful to model the encoding of clinical cognition into narrative using discourse-based theory. This non-linearity, whether expressed by a study of story sequencing or cognition, is theorized by *Kay and Purves* to be a central component of the clinical narrative. Specifically, while the medical record is primarily comprised of a narrative (such that the narrative is representative of genres of clinical writing [38, 39]), discourse is produced and integrated between the physician and the patient as authors in multiple instances across the narrative or more generally, the series of narratives which make up the clinical encounter.

Therefore, we can conclude that the clinical narrative is composed of individual encounter narratives, and is contextualized as a non-linear process (a metanarrative) integrating the fabula and sjuzhet across multiple encounters. It is therefore useful to characterize the sjuzhet of such a meta-narrative through the lens of seriality or sequencing. Between encounters, there are a number of events that interrupt the sequence, as well as there is an inherent non-linearity to the collection of clinical temporal data. For example, while nursing may collect vitals on the wards every four hours, it is not always recorded into the medical record immediately following collection. Such a delay inherently places an incongruence between clinical event time and recorded event time, resulting in a non-linear sequence between these events in the narrative of the patient’s medical record. Within the clinical note, non-linearity is observed when the note is constructed, by disparate structures of writing being used to sequence events. From the SOAP/APSO note [40] to the standard history/physical/review of systems sections to formal clinical templates, clinical events are not communicated in a strict linear fashion, nor are they made explicitly temporally congruent with clinical event time.

In reference to the forms of a serial narrative, *Sherzer* [41] and *O’Sullivan* [42] suggest elements with which such a meta-narrative as the clinical encounter can be modeled using the examples of a metanarrative (using the exemplar of Claude Simon’s *Triptyque* [43]). Outside the genre of the *Nouveau Roman*, a more recent example of meta-narrative can be found in David Foster Wallace’s *Infinite Jest* [44], the narrative disentanglement of which reveals a non-linear meta-narrative structure. [45] *Sherzer* suggests that non-linearity, discontinuity and fragmentation, while *O’Sullivan* suggests that iteration, multiplicity, momentum, world-building, personnel and design are the properties that characterize and organize text.

Combining these two suggested sets of elements, the clinical meta-narrative can be characterized by a nonlinear connection of events which are both separated by discontinuity and internally fragmented. Furthermore, these events are often repetitive (such as multiple instances of history-taking) and display differing and evolving characters and context (such as multiple visits for a child with differing parents or grandparents attending). We can therefore conclude that the meta-system which we call the clinical narrative is best analogized to a series of generative systems. We can therefore articulate this notion of a serial encounter-driven meta-narrative through the nature of events within the encounter at the level of the generative system, rather than the structure system of the encounter or the meta-system of the narrative. This terminology is taken from the observations of complexity by *Klir* [46], in which the observation of a set of things and relations are encapsulated by a system (distinguished by the events within a clinical encounter) and provides an ontological separation between generative, structure and meta-system systems. The meta-system refers to the system distinguished by the entirety of the clinical narrative (its objects being the clinical encounters), and the structure system the clinical encounters themselves (their objects being the systems of clinical tasks therein), and the generative system the individual clinical events within the encounter (e.g. the task of physical examination including the events of taking a blood pressure, weight, height…).

A generative system is one which is support-invariant (in that it is independent from the overarching structure in some fashion, typically seen in time-invariant systems), which in this case is indicative of the clinical invariance found in the individual encounter. A structural system is one which generative systems interact, which in this case is for diagnostic purpose, as while clinically invariant, their information lends to an overall diagnosis across encounters (as in the nonemergency setting, a diagnosis is often not made until after the information from first encounter has been parsed). A meta-system system is a system in which the systems encapsulated by the meta-system is allowed to change within the support set of the metasystem (for example, a time-varying finite state machine). A clinical example of a meta-system would be the primary care practice. For example, a child who comes in to a regular visit with their pediatrician, and is sent for further evaluation by a specialist who in turn sends out for imaging from a radiologist, prior to the child returning to their pediatrician for the followup encounter, parsing both new clinical information gained, including the report from the specialist (who in turn parses the report from the radiologist), and rendering a diagnosis. In this example, the clinical work is invariant between each physician, separating each clinical task and encounter within the patient’s narrative. Such a patient’s clinical narrative can therefore be thought of as complex, because complexity is found in such systems which are a system of systems, across hierarchies. [47]

### Linguistic Analysis

Linguistic analysis, whether computationally or manually performed in the domain of clinical medicine has not addressed the nature of the clinical narrative, or the nature of narrative text. Instead, this analysis typically focuses on character-encoded text as information (a concept suggested by Shannon in his seminal treatise [48]) or dialogic methods. This is exemplified by efforts to mathematically model abstractions of clinical text [49, 50], which facilitate structure for both summarization and relation classification. These analyses have failed to investigate the clinical narrative, but rather impose a structure that is in contrast to that of narrative. We can similarly find this from earlier efforts to extract linguistic information from text [51], which demonstrate syntagmic or propositional fidelity of information, but do not address more granular linguistic constructs such as discourse. [52] Efforts to examine narrative constructs within medicine have focused on an anthropological viewpoint of physicians and patients [53–55] or a patient-centered cultural viewpoint [56, 57], which in turn has led to suggestions of discourse based on a cultural framing. [58, 59] The suggestion, therefore, has been that the clinical narrative, separate from the clinical narrative *text* is representative of “reasoning as a socially mediated activity” rather than addressed as the entanglement between cognition and language present at the heart of the information used for the documentation task. [60]

By constructing a meta-model of information within the medical record, we account for a narrative structure to said data. The model must accurately articulate the fabula elements which equivocate to diagnostic narrative énoncé elements, integrated by a sjuzhet which respects the connections between both fabula elements and their constituents. While we have described the model diagrammed in Figure 2 in philosophical and linguistic terms, its implementation is fundamentally tied to the lexicalization of clinical concepts. As a source of data to validate the model, we therefore need a source of clinical text written by a physician. To facilitate (external) semantic evaluation by the non-physician reader, this note should be written as an expository narrative, rather than an argumentative record of scientific observation (a chronicle). [61] While both are acceptable forms of clinical narratives, the latter is often paraphrasic and terse to the point of unintelligibility by the non-physician (particularly in reading deictic clinical information). With such a requirement in mind, the University of Rochester’s combined medicine and psychiatry service [62] textbook by *Morgan and Engel* entitled “The Clinical Approach to the Patient” [63] contains a narrative verbatim interview and case write-up of a patient. In particular, the diagnoses in this case are laid bare in a section illustrating the context of an enumerated list of diagnoses. Clinically these diagnoses are the fabula elements of the clinical encounter. This note, while not current, presents a clear picture of the clinical encounter, including information often excluded from notes [64, 65] gleaned from the interview transcript. This clarity facilitates explication ahead of clinical semantic problems which arise from a lack of clinical expertise by the non-physician reader, and offers as robust of a viewpoint on the clinical encounter as we can get without a video record during a retrospective case review.

In producing the analyses which follow, the lead author (DC) manually performed the linguistic analysis and subsequent textual annotation, validated by both linguistic (MD) and clinical experts (SMD). MBR). This is necessitated as noted above, due to the fact that existing automated technology insufficiently addresses the linguistic concepts applied to clinical medicine. Additionally, as this analysis is not performed at scale, a computational approach does not provide value beyond introducing domain-related error (which would necessitate manual correction).

### Data Source

The case note from *Morgan and Engel* describes the case of “a 52-year-old married woman was admitted to the hospital from another city for an evaluation of symptoms of dyspnea, orthopnea, edema, and rapid heart action. She had recently suffered from right-sided weakness.” [66] The patient has the following list of diagnoses, listed with corresponding Unified Medical Language System (UMLS) Concept Unique Identifiers (CUIs) in square brackets:

1. Chronic rheumatic heart disease [C0175708] with predominant mitral stenosis [C0026269] and slight mitral regurgitation [C0026266], Functional Class III. [C1882086]
2. Atrial fibrillation. [C0004238]
3. Congestive heart failure (by history). [C0018802]
4. Right arm paresis [C0856328], secondary to left middle cerebral artery embolus [C4543471] (6 weeks ago).
5. Depression [C0011570], occult [C0205262].

While occult depression is noted in the prognosis, this diagnosis stems from the patient interview, and is not noted in the diagnosis section which contains the medical context of the noted coronary and circulatory system diagnoses. As such, we can exclude depression from our list of diagnoses with which to analyze.

### Annotation

Each of the aforementioned diagnoses are described narratively within the *Diagnosis* section of the note. We can conclude that the fabula elements of the diagnosis section are therefore the four diagnostic problems, excluding occult depression. The topic sentences of each of the paragraphs map to each of the enumerated diagnoses, with sentences clustering together as multiple paragraphs detail two diagnoses. We can use these as approximate fabula elements, albeit with the separation of “Chronic rheumatic heart disease / with predominant mitral stenosis and slight mitral regurgitation, / Functional Class III.” as the diagnosis is representative of three items across three paragraphs, unlike “Right arm paresis, secondary to left middle cerebral artery embolus (6 weeks ago).” which is within a single paragraph. Each topic sentence is presented as segmented into elementary discourse units.

1. Chronic rheumatic heart disease with mitral stenosis and slight mitral regurgitation, Functional Class III
  a. Chronic rheumatic heart disease [The patient, who is 52 years old with possible rheumatic fever at age 8,]^1*A*^ [has had an intermittent cardiac irregularity for 25 years and the gradual progression of dyspnea, orthopnea, and dependent edema over 4 to 5 years.]^1*B*^
  b. with mitral stenosis and slight mitral regurgitation [The cardiac findings are classically those of mitral stenosis and mitral regurgitation,]^2*A*^ [the predominant lesion being mitral stenosis.]^2*B*^
  c. Functional Class III [The gradually developing dyspnea, orthopnea, and more persistent ankle edema 4 to 5 years ago]^3*A*^ [marks the beginning of cardiac decompensation.]^3*B*^
2. Atrial fibrillation
  a. [The history of periodic irregular heart action since 1945 is compatible with paroxysmal atrial fibrillation,]^4*A*^ [which is not uncommon in mitral stenosis,]^4*B*^ [but these symptoms could also have been due to intermittent premature beats.]^4*C*^
  b. [The episode of cardiac irregularity and transient pain in August, 1967,]^5*A*^ [suggests the sudden onset of atrial fibrillation with a rapid ventricular rate]^5*B*^ [resulting in decreased cardiac output and coronary insufficiency.]^5*C*^
3. Congestive heart failure [The symptoms of congestive heart failure began 4 or 5 years ago,]^6*A*^ [but may also have been present during the three pregnancies in 1946, 1951, and 1957,]^6*B*^ [precipitated by the increased blood volume of pregnancy.]^6*C*^
4. Right-arm paresis, secondary to left middle cerebral artery embolus [The sudden right hemiparesis, aphasia, and right homonymous hemianopsia of December 11,1967,]^7*A*^ [suggest an arterial obstruction]^7*B*^ [in the distribution of the left middle cerebral artery.]^7*C*^

### Propositional Perspective

In the biomedical/clinical domain, clinical concepts and their semantic relations have been described by the Semantic Knowledge Representation Project (SemRep) [67], specifically acting as a gold standard given knowledge sourced in literature. [68] SemRep uses the UMLS as the basis for its semantic structures, providing an access point to semantic relations and predication through a separate database, SemMedDB. [69] The UMLS is a system which contains a meta-thesaurus connecting clinical vocabularies, terminologies and standards together as CUIs via both hierarchical relations and a semantic network.

As noted by *Lemieux and Bordage*, the propositional perspective is “an appropriate selection of relevant clinical findings”. [4] In the model, fabula elements contain individual clinical signs and symptoms. Fundamentally, the term *relevant* implies a semantic structure, and *appropriate* can be taken to mean those which would be within the clinical vocabulary. As indicated by the folkloric examples, the fabula elements encapsulate non-linear semantic information both within and between énoncés. Using the UMLS as a basis for the clinical vocabulary, we can extract CUIs within the énoncés making up a fabula element, and generate a network given known hierarchical and semantic relations. These edges are reflective of the semantic predication encoded within the UMLS. [68]

For example, in the topic sentence la, such a network is detailed in Figure 4. In this sentence six UMLS CUIs can be identified: patient, C0030705; rheumatic fever, C0035436; cardiac irregularity, C1401284; dyspnea, C0013404; orthopnea, C0085619; dependent edema, C0235437. While the concept of “year”, and “age” can be found within the UMLS (whether as age-years C1510829, year C0439234, or age C0001779), the concept of “52 years of age”, “at age 8”, “25 years” or “4 to 5 years” are qualifiers of the patient or rheumatic fever. As such, they can be discarded when selecting core concepts within the text which are semantically connected to others (rather than a concept that is fundamentally a qualification). In addition, these qualifiers provide semantic information for the edges of the graph, enabling the qualification of the relationship between most concepts: PRECEDES, which has a Term Unique Identifier of T138. The other semantic relationship in this sentence is the relationship between patient and rheumatic fever (HAS MANIFESTATION, T150).

**Figure 4.**
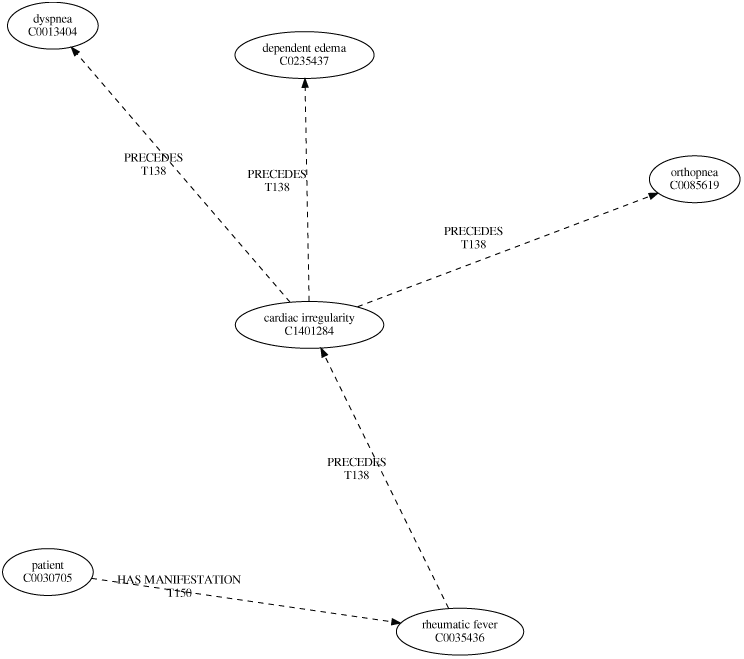
Propositional Perspective of Concepts. For the sentence: *The [patient]*^*C*0030705^, *who is 52 years old with possible [rheumatic fever]*^*C*0035436^ *at age 8, has had an intermittent [cardiac irregularity]*^*C*1401284^ *for 25 years and the gradual progression of [dyspnea]*^*C*0013404^, *[orthopnea]*^*C*0085619^, *and [dependent edema]*^*C*0235437^ *over 4 to 5 years*. Concepts are paired with their Concept Unique Identifier, while relations are paired with their Term Unique Identifier from the UMLS.

### Structural Semantic Perspective

*Lemieux and Bordage* describe the structural semantic perspective as “an abstraction of [appropriately selected relevant clinical] findings into pertinent networks of formal qualities.” [4] Such a network was previously developed by *Lemieux and Bordage* [6], building upon the symbology of *Propp*. [70, 71] This domain-specific structure relies on symptoms and signs as constituent units, and a morphological characterization of the reasoning operations and competencies of the physician. While this is useful in articulating the actions taken during a transcript of patient-provider discourse, it. lacks applicability to the information encoded by said reasoning in the discourse of the clinical note. For example, the competency of “clarifying and ambiguous or imprecise clinical cue” (?) can be articulated by an interrogative statement present, in patient provider discourse, or an abstraction of a series of reasoning statements in the subsequent, written note. In such an example, the coding schema would be applied at the fabula element level, rather than that of the individual discourse unit(énoncé). Fundamentally the ambiguity resulting in examples of the latter, in which a reasoning task or competency spans multiple elementary units of written discourse, is a clear example of the lack of applicability of the coding schema. As the constituent units of the information model (signs and symptoms) are contained within discourse units (the diagnostic énoncé), an abstract semantic network that connects said units is necessary. Rhetorical Structure Theory (RST) has been noted as such a theoretical framework.

With this in mind, we can apply RST to topic sentence la in Figure 5, connecting the discourse units (énoncés) detected. Specifically, for the topic sentence, the semantic relation of BACKGROUND connects the description of patient with possible history of rheumatic fever to symptoms (cardiac irregularity, gradual progression of dypsnea, orthopnea and dependent edema). In this ease, the nucleus of the sentence (the presence of symptoms of rheumatic fever) is connected to by the satellite describing a possible prior history of rheumatic fever. This provides a network relation which suggests that the symptoms are related to the history.

**Figure 5.**
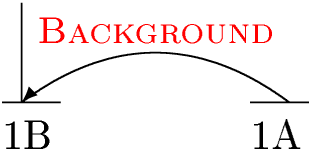
Structural Semantic Perspective of Concepts. For the sentence *[The patient, who is 52 years old with possible rheumatic fever at age 8,]*^*A*^ *[has had an intermittent cardiac irregularity for 25 years and the gradual progression of dyspnea, orthopnea, and dependent edema over 4 to 5 years*.]^*B*^

### Integration (Calibration) of Perspectives

Integrating the propositional and structural semantic perspectives completes the information model for the clinician diagnosing a patient given signs and symptoms, as well as a known disease. Using *Klir’s* hierachy [46], given that the source system of signs and systems is encapsulated within a data system of lexical items, and is abstracted into a generative system by each of the perspectives, a meta-system represents the system articulated by the diagram in Figure 2. At the encounter-level, the model is integrating two systems of concepts connected by semantic relations (producing a structure system) at different hierarchical levels (the UMLS CUI and énoncé). Both in terms of the structure system and the overarching meta-system, the model which integrates the perspectives is a complex network.

Within the dual-process theory based universal model of diagnosis proposed by *Croskerry* [7], calibration is the last stage prior to achieving the stage of diagnosis. Calibration is the post-processing of system I and system II output, such that they interact to produce a diagnosis. In the clinical semiotic sense, this is the production of closure from the identification of signs and symptoms, and disease. Similarly, in the meta-model of the clinical narrative, the integration of selected relevant clinical findings, and their abstraction into a network relating these findings together produces semiotic closure. This closure is present within the resulting network diagram, which connects diagnostic énoncés by the underlying propositional structure of UMLS CUIs, as well as the rhetorical structure of the medical discourse.

Figure 6 provides an example of this integration for topic sentence la. Closure, as represented between the énoncés is facilitated by the PRECEDES relationship between rheumatic fever and cardiac irregularity, in parallel to the BACKGROUND from the rhetorical structure. This provides a clear articulation that temporal precedence is what provides meaning to the background connection between the patient’s history of rheumatic fever and expression of symptoms.

**Figure 6.**
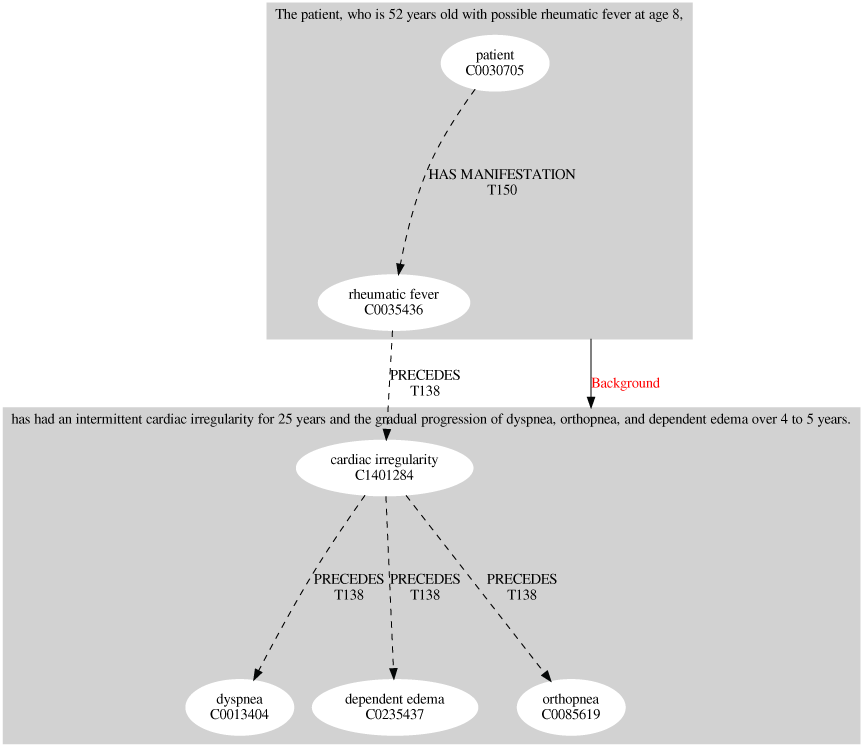
Propositional and Structural Semantic Perspectives Concept Integration. For topic sentence 1a, integrating Figures 4 and 5.

## Results

In articulating the perspectives, topic sentence la has been used as an example, producing Figures 4, 5, and 6. Each of the remaining topic sentences (lb through 4), are described in Figures 10 through 21. Additionally, integration diagrams are described in Figures 22 through 27. Each of these figures describes a component of the overarching fabula element of the diagnostic report, an individual diagnosis (or component therein) a piece. The collection of these figures together into a single model is the complex model of the clinical narrative of the diagnostic report. Figure 8 details the model of the clinical narrative, combining the integration diagrams, and demonstrating closure between topic sentences. This model uses the topic sentences as fabula elements, and therefore integrates the sub-fabula connectives as a component of the sjuzhet from a propositional perspective. The sjuzhet modeled as structural semantic connectives requires additional information than the relations between sub-fabula elements can provide. This additional information is the ordering and semantic relations between the topic sentences as they appear in the text:

**Figure 7.**
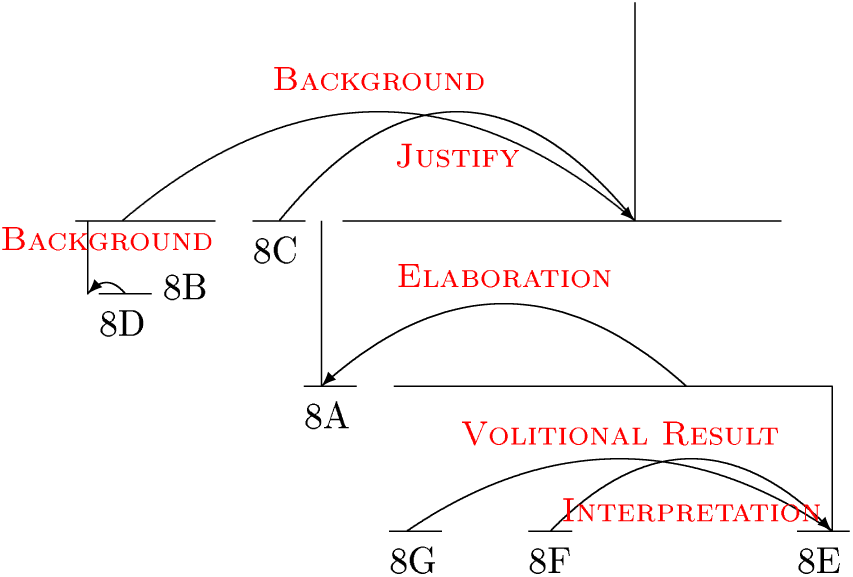
Rhetorical Structure Model of Sjuzhet from Topic Sentence Order in Text. For topic sentences 1a through 4 as ordered in the narrative.

**Figure 8.**
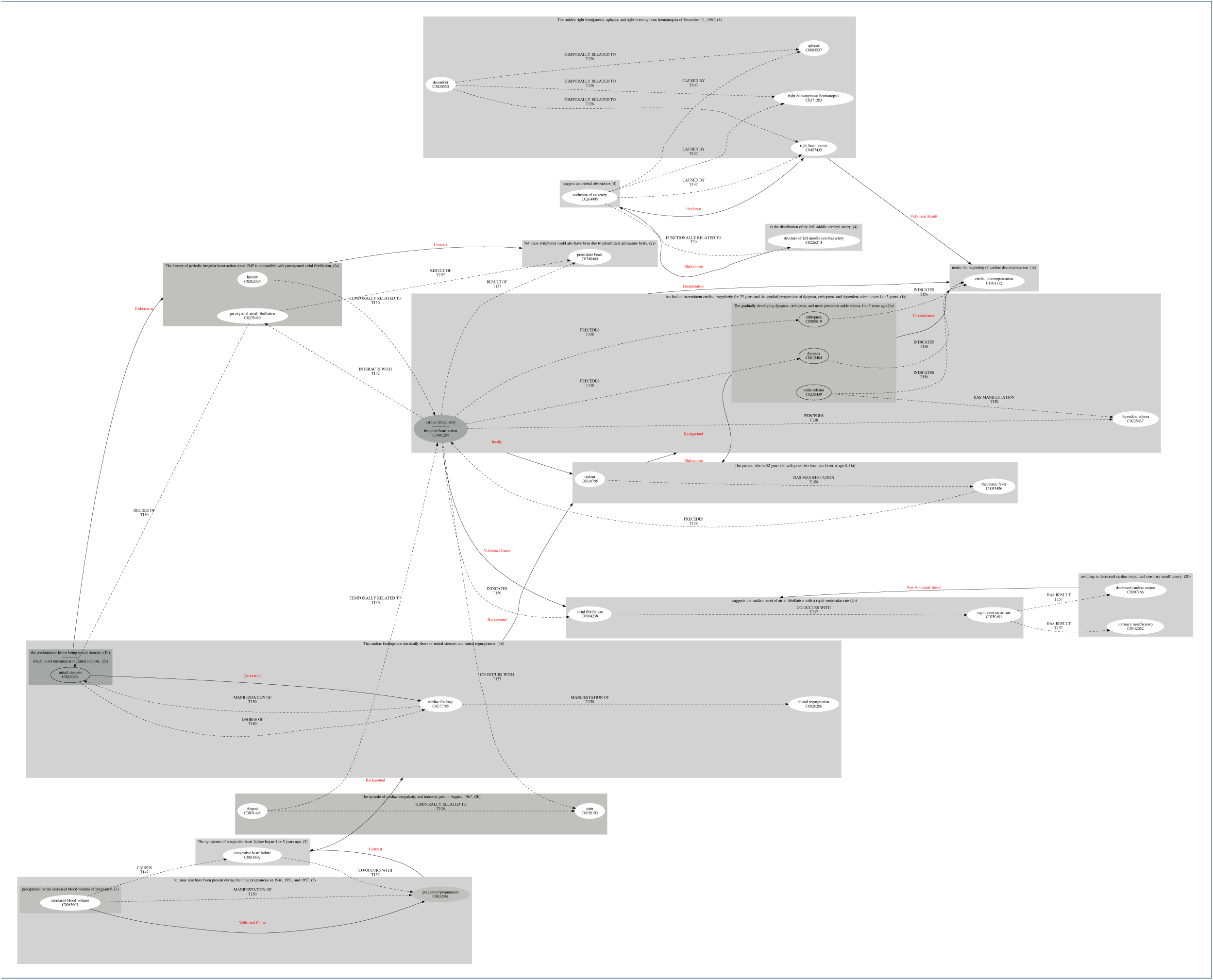
Clinical Narrative Meta-Model as a Network. For topic sentences 1a through 4, integrating diagnostic énoncés and UMLS concepts. Diagnostic énoncés are outlined in lightgrey and are followed by parenthetical topic sentence identifiers. Diagnostic énoncé cannot intersect. Concepts which span two énoncés are outlined in grey. Concepts which span three énoncés are outlined in grey65.

**Figure 9.**
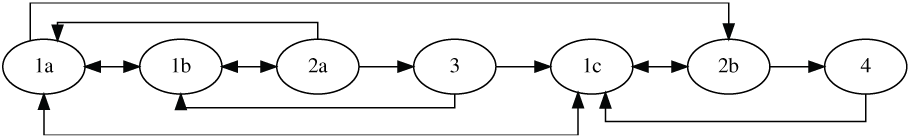
Narrative Sequence-Semantics of Topic Sentences. Consistent with the pictorial representation of narrative structure developed by *Trabasso* [36], the sequence, propositional and structural semantic relations present between topic sentences 1a through 4.

**Figure 10.**
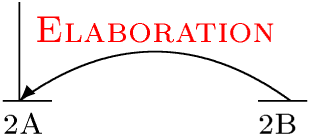
Structural Semantic Perspective of Concepts. For the sentence *[The cardiac findings are classically those of mitral stenosis and mitral regurgitation,]* ^*A*^ *[the predominant lesion being mitral stenosis*.]^*B*^

**Figure 11.**
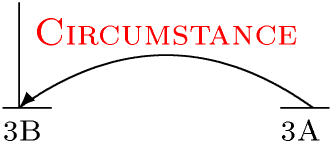
Structural Semantic Perspective of Concepts. For the sentence *[The gradually developing dyspnea, orthopnea, and more persistent ankle edema 4 to 5 years ago]*^*A*^ *[marks the beginning of cardiac decompensation*.]^*B*^

**Figure 12.**
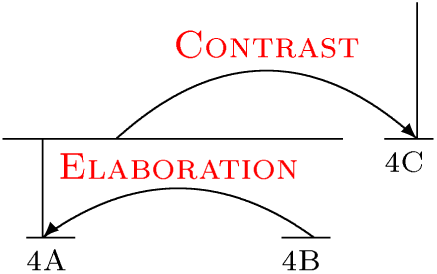
Structural Semantic Perspective of Concepts. For the sentence *[The history of periodic irregular heart action since 1945 is compatible with paroxysmal atrial fibrillation,]*^*A*^ *[which is not uncommon in mitral stenosis,]*^*B*^ *[but these symptoms could also have been due to intermittent premature beats*.]^*C*^

**Figure 13.**
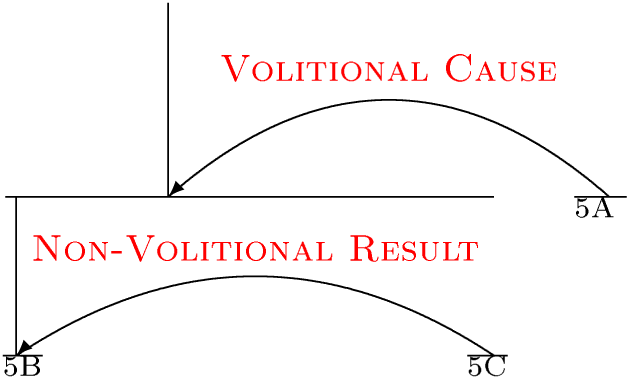
Structural Semantic Perspective of Concepts. For the sentence *[The episode of cardiac irregularity and transient pain in August, 1967,]*^*A*^ *[suggests the sudden onset of atrial fibrillation with a rapid ventricular rate]*^*B*^ *[resulting in decreased cardiac output and coronary insufficiency*.]^*C*^

**Figure 14.**
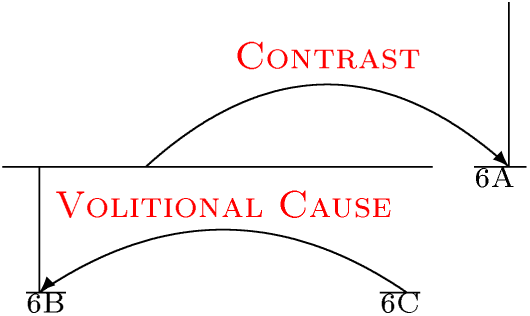
Structural Semantic Perspective of Concepts. For the sentence *[The symptoms of congestive heart failure began 4 or 5 years ago,]*^*A*^ *[but may also have been present during the three pregnancies in 1946, 1951, and 1957,]*^*B*^ *[precipitated by the increased blood volume of pregnancy*.]^*C*^

**Figure 15.**
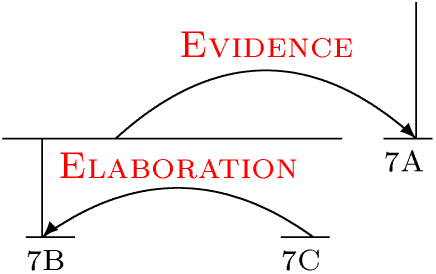
Structural Semantic Perspective of Concepts. For the sentence *[The sudden right hemiparesis, aphasia, and right homonymous hemianopsia of December 11, 1967,]*^*A*^ *[suggest an arterial obstruction]*^*B*^ *[in the distribution of the left middle cerebral artery*.]^*C*^

**Figure 16.**
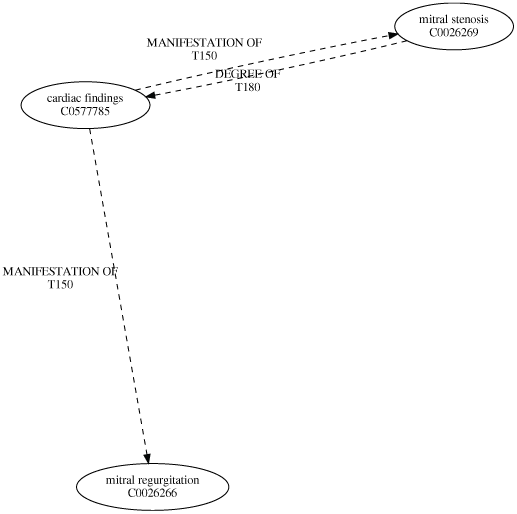
Propositional Perspective of Concepts. For the sentence: *The [cardiac findings]*^*C*0577785^ *are classically those of [mitral stenosis]*^*C*0026269^ *and [mitral regurgitation]*^*C*0026266^, *the predominant lesion being [mitral stenosis]*^*C*0026269^. Concepts are paired with their Concept Unique Identifier, while relations are paired with their Term Unique Identifier from the UMLS.

**Figure 17.**
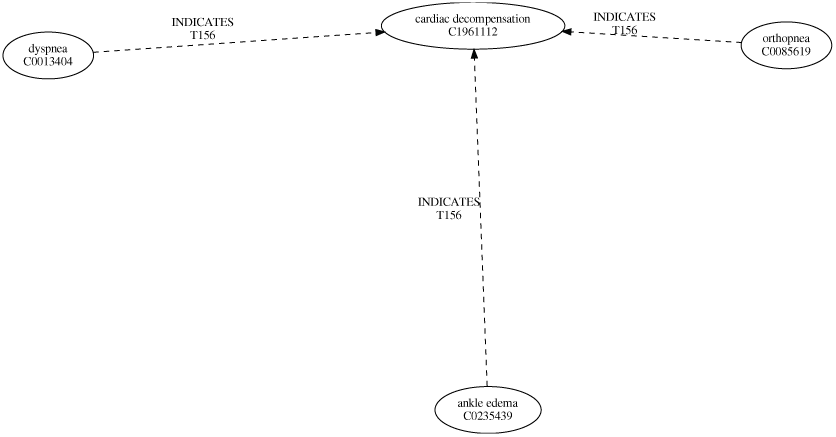
Propositional Perspective of Concepts. For the sentence: *The gradually developing [dyspnea]*^*C*0013404^, *[orthopnea]*^*C*0085619^, *and more persistent [ankle edema]*^*C*0235439^ *4 to 5 years ago marks the beginning of [cardiac decompensation]*^*C*1961112^. Concepts are paired with their Concept Unique Identifier, while relations are paired with their Term Unique Identifier from the UMLS.

**Figure 18.**
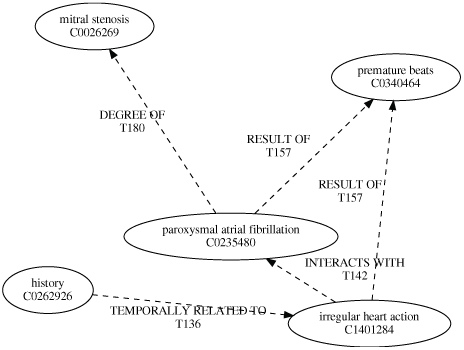
Propositional Perspective of Concepts. For the sentence: *The [history]*^*C*0262926^ *of periodic [irregular heart action]*^*C*1401284^ *since 1945 is compatible with [paroxysmal atrial fibrillation]*^*C*0235430^, *which is not uncommon in [mitral stenosis]*^*C*0026269^, *but these symptoms* ^*C*1401234^ *since could also have been due to intermittent [premature beats]*^*C*0340464^. Concepts are paired with their Concept Unique Identifier, while relations are paired with their Term Unique Identifier from the UMLS.

**Figure 19.**
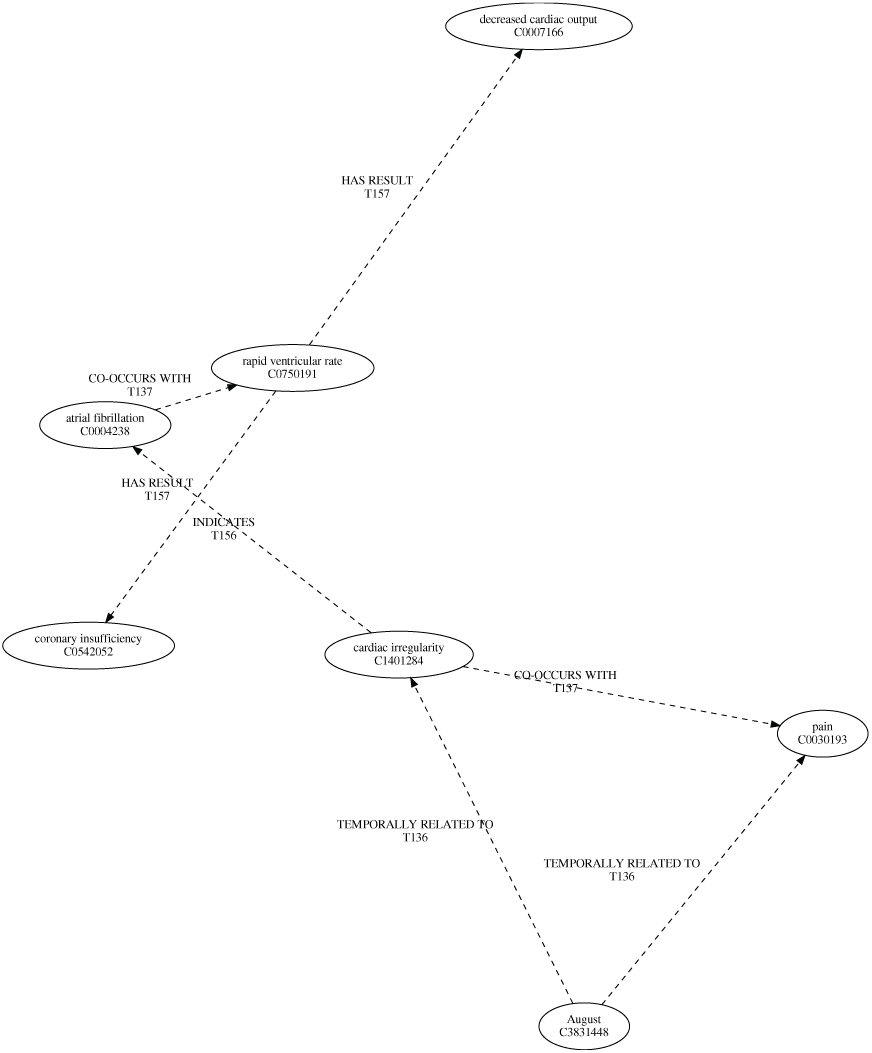
Propositional Perspective of Concepts. For the sentence: *The episode of [cardiac irregularity]*^*C*1401284^ *and transient [pain]*^*C*00030193^ *in [August]*^*C*03831448^, *1967, suggests the sudden onset of [atrial fibrillation]*^*C*0004238^ *with a [rapid ventricular rate]*^*C*0750191^ *resulting in [decreased cardiac output]*^*C*0007166^ *and [coronary insufficiency]*^*C*0542052^. Concepts are paired with their Concept Unique Identifier, while relations are paired with their Term Unique Identifier from the UMLS.

**Figure 20.**
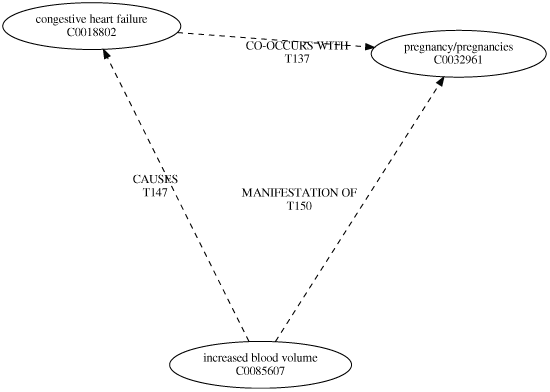
Propositional Perspective of Concepts. For the sentence: *The symptoms of [congestive heart failure]*^*C*0018802^ *began 4 or 5 years ago, but may also have been present during the three [pregnancies]*^*C*0032961^ *in 1946, 1951, and 1957, precipitated by the [increased blood volume]*^*C*0085607^ *of [pregnancy]*^*C*0032961^. Concepts are paired with their Concept Unique Identifier, while relations are paired with their Term Unique Identifier from the UMLS.

**Figure 21.**
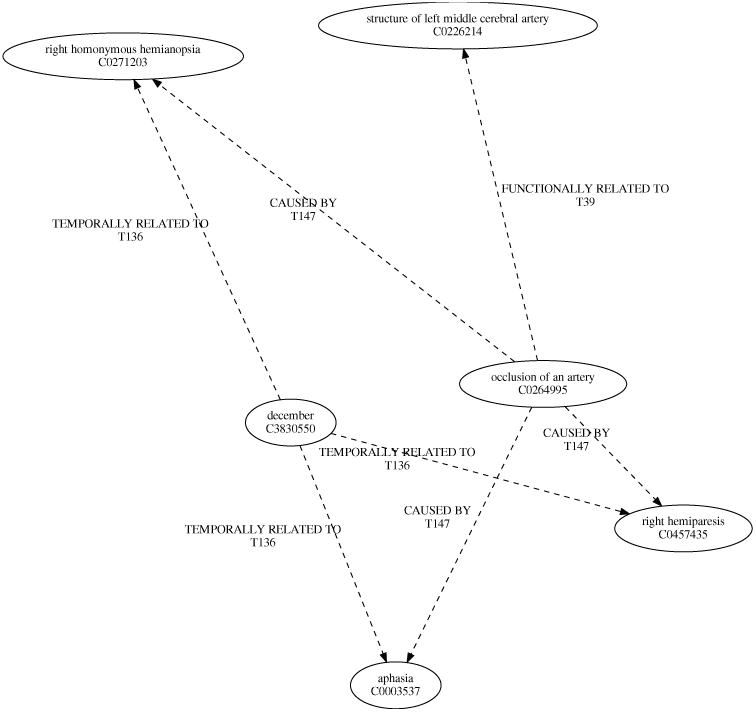
Propositional Perspective of Concepts. For the sentence: *The sudden [right hemiparesis]*^*C*0457435^, *[aphasia]*^*C*0003537^, *and [right homonymous hemianopsia]*^*C*0271203^ *of [December]*^*C*3830550^ *11, 1967, suggest an [arterial obstruction]*^*C*0264995^ *in the [distribution of the left middle cerebral artery*.]^*C*0226214^ Concepts are paired with their Concept Unique Identifier, while relations are paired with their Term Unique Identifier from the UMLS.

**Figure 22.**
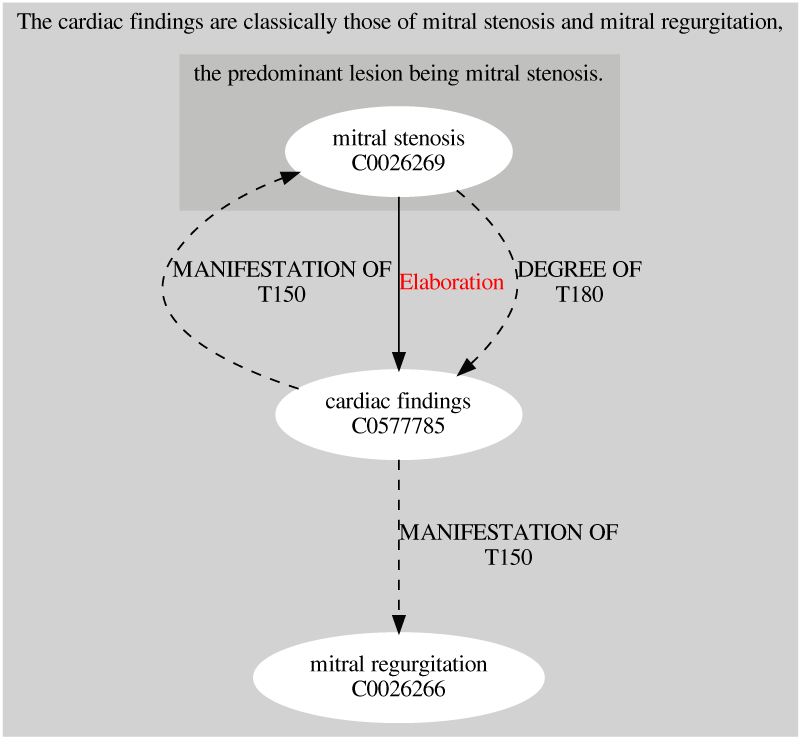
Propositional and Structural Semantic Perspectives Concept Integration. For topic sentence 1b, integrating Figures 10 and 16.

[Chronic rheumatic heart disease (topic sentence la)]^8*A*^ [With mitral stenosis and slight mitral regurgitation (topic sentence lb)]^8*B*^ [Atrial fibrillation (topic sentence 2a)]^8*C*^ [Congestive heart failure (topic sentence 3)]^8*D*^ [Functional Class III (topic sentence lc)]^8*E*^ [Atrial fibrillation (topic sentence 2b)]^8*F*^ [Right arm paresis, secondary to left middle cerebral artery embolus (topic sentence 4)]^8*G*^

Closure outside of the integration diagrams via RST is facilitated by the connection of fabula elements, detailed in Figure 7. This provides clarification for the semantic relations between specific concepts as key components of the connected fabula elements. These rhetorical relations connect the right hemiparesis, aphasia and homonymous hemianopsia with cardiac decompensation; a volitional cause/rcsult relationship. The presentation of cardiac findings determined by mitrial stenosis and regurgitation are rhetorically related as background information for congestive heart failure. The August 1967 episode of cardiac irregularity is rhetorically related as an instance of background for the 25 year history of intermittent cardiac irregularity. The marked instance of cardiac decompensation is presented as interpretive of the past cardiac irregularity. The gradual progression of dyspnea (as well as orthopnea and dependent edema) is seen to elaborate on the patient’s state. In addition, cardiac irregularity is seen to justify the same symptoms, which is further suggestive of their relation to cardiac decompensation and subsequent congestive heart failure.

Closure outside of the integration diagrams via UMLS semantic relations is facilitated by a UMLS concept being common to multiple énoncés. The concepts of cardiac irregularity/irregular heart action (C1401284), and mitral stenosis (C0026269) span three énoncés. The concepts of pregnancy (C0032961), ankle edema (C0235439), orthopnea (C0085619), and dyspnea (C0013404) span two énoncés. This commonality results in three sets of énoncés completely overlapping:

1. “the predominant lesion being mitral stenosis.” *and* “which is not uncommon in mitral stenosis,”
2. “The gradually developing dyspnea, orthopnea, and more persistent ankle edema 4 to 5 years ago” *and* “has had an intermittent cardiac irregularity for 25 years and the gradual progression of dyspnea, orthopnea, and dependent edema over 4 to 5 years.”
3. “but may also have been present during the three pregnancies in 1946, 1951, and 1957,” *and* “precipitated by the increased blood volume of pregnancy.”

This closure is primarily facilitated through the concepts of cardiac irregularity/irregular heart action. It connects the cardiac irregularity/irregular heart action which is a symptom of chronic rheumatic heart disease, as well as a component of atrial fibrillation. Fundamentally, this provides a semantic connection between observed cardiac irregularity and heart disease, be it rheumatic or atrial. Mitral stenosis does not contribute to diagnostic closure, as it only connects énoncés within a single diagnosis (that of topic sentence lb). Secondarily, the trio of ankle edema, orthopnea and dyspnea facilitate closure between topic sentences la and 1c. This closure expected given the narrative, and provides a semantic link via the trio of concepts to the qualification of chronic rheumatic heart disease as being of functional class III.

## Discussion

The network model of clinical narrative provides a mechanism with which we can frame a complex model of medical information. The complexity is wrought by the non-linearity inherent in the meta-model’s construction. Mechanistically, for the narrative, diagramming the sequence of diagnoses represented by the sequence of topic sentences (and therefore paragraphs) provides a means with which this non-linearity can be simplified from Figure 8. This diagram is patterned after the recursive causal transition network diagrams of *Trabasso*, which provide both a diagram as complex as that of Figure 8 (in the case of *The Father, His Son and Their Donkey* story [72]), and a simplified structure representative of an abstraction of story events. Similarly to Proppian symbology, the recursive causal transition network diagram provides a representation of both the fabula and the sjuzhet. For the clinical case exemplar, Figure 9 describes such a network, abstracting the network in Figure 8 into the topic sentences and the sjuzhet as a combination of sequence, propositional and structural semantic relations.

Beyond complexity, the meta-model frames a lens to examine clinical information. In framing the nature of reasoning and information in medicine, *Blois* suggests that classifying disease in medicine is a vertical reasoning task, connecting hierarchies of information from the patient as a whole down to the molecules and atoms representative of chemistry and physics. For example, in the case of Wilson’s disease, the patient’s malaise, schizophreniform disorders and labile affect are linked to the physiologic intention tremor, dysarthria, dystonia and Babinski sign, which in turn are linked through organs, cells and biochemistry to the clinical chemistry of aminoaciduria, decreased scrum copper and increased urinary copper. This inter-hierarchical model of disease classification informs the end product of diagnosis, in that as an act of meaning the signs and symptoms of the previous sentence are linked to Wilson’s disease. Wilson’s disease is classified as a metabolic disorder of mineral metabolism by the International Classification of Diseases, Tenth Revision, Clinical Modification (ICD-10-CM). Medical information to diagnose Wilson’s disease, therefore, is complex in its presentation as signs and symptoms. Integrating this information through the task of clinical reasoning therefore requires a complex meta-model. The relations drawn between each of the hierarchical sources of information in integrating this complex model in the diagnostic act are those which provide clinical semiotic closure, linking signs and symptoms to the disease through the assignment meaning.

The nature of clinical information can therefore be characterized as relative to an entity beyond that of the perspectives from which it is represented. Whether relative to the normal as described by *Canguilhem* [73] or to the reference point of clinical nosology (such as that of ICD-10-CM), the result is the integration of multiple systems. Consider the investigation of fever, in which a causal agent renders a nosological difference (such as a bacteriological investigation which changes a diagnosed influenza to typhoid fever). In this case, it is the nature of signs and symptoms such that they are used to create a meaning assignment to an articulated bacteriological nosology of disease. *Crookshank* [74] articulates that the clinical semiotic closure is precipitated by the linguistic entity at the core of the act of diagnosis. In the course of a clinical narrative, be it historical or in an acute setting, *Crookshank* suggests that the act of diagnosis is tied to pronunciation of a disease by name.

As such, the interrogation of the language used to detail signs and symptoms and name the disease in the course of diagnosis, is a key component of articulating the nature of information in the clinical narrative. The meta-model provides a viewpoint which one can investigate the relationship between information in medicine and grammar; betwixt which is the clinical sub-language. It is therefore meaningful that as an interstitial force the clinical sub-language has its roots in a Galenic approach to medicine. *Sluiter* [75] describes the necessity of the grammatical education required by Galen in the formative years of a physician’s career as propaedeutic, in that it contributes to the framing of description in medicine. *Buchanan* [76] furthers Galen’s antiquarian viewpoint and links grammar and the other liberal arts of the trivium and quadrivium to medicine through clinical semiotics. In both a Galenic and semiotic approach, understanding of the grammatical morphology of the clinical sub-language (the interplay of syntax and semantics) supports a clearer picture of the nature of medical description and more accurate diagnosis within the context of both clinical relativism and measurement. In his seminal treatise on the nature of relative pathological measurement, *Canguilhem* clarifies such a role of language by using the concept of information theory as a mechanism to encode medical action; that which is precipitated by clinical reasoning and scientific entailment at pointof-care. The sub-language of medicine, therefore, requires attention to the morphology of both the clinical concepts and their measurement, such as both of these tasks result in diagnosis through the assignment of semantic relations. Formally, sub-language analysis offers a grammatical characterization and structure problem akin to that of diagnosis. [77]

To solve the characterization and structure problem is to resolve the clarity with which medicine can be represented by language in discursive form. Mechanisms of description from a propositional perspective are insufficient to encapsulate the vocabulary of clinical practice. This insufficiency necessitates a structural semantic perspective to clarify relationships at a granular level of language rather than for the propositional concept-element. A clear example of this is pain, where the articulation of the nature of pain is not included in ontological strictures given its sign (symptom) status. Yet this leads to diagnostic and clinical judgement uncertainty for a greater proportion of generalist care than specialist medicine. [78]

A structured model such as that of the meta-model affords a systematic approach to the analysis of both linguistic forms and concepts derived from clinical reasoning. This approach clarifies an uncertainty of the clinical sub-language, as well as supports the underlying semiotic approach to diagnosis through the integration of clinical reasoning and the medical information model. *Meyer and Singh* [10], describing the path to diagnostic excellence through calibration detail a potential application for the model. The descriptive capacity of language can be harnessed to provide feedback during the clinical reasoning process via semiotic explanations. Specifically, such decision support could intervene at the calibration stage of clinical reasoning, such that physicians are supported in managing the complexity of diagnosis due to the patient and situational factors of information integration as they are entered into the electronic medical record. Such decision support would inherently function on information provided by the task of documentation by the physician, and therefore has an educational purpose in supporting the fifth entrustable professional activity developed by the Association of American Medical Colleges, to “document a clinical encounter in the patient record”.

## Conclusions

In conclusion, this paper proposes a complex model of diagnostic information. The model is demonstrated through the use of narrative information, both in terms of text and story. The model meets *Klir’s* definition of a meta-model, in that it is a complex connective of a multi-hierarchical and structured system of generative systems. This meta-system addresses semantic information problems at the heart of diagnosis, first articulated by *Allbutt*, explicated by both clinical semiotics and reasoning as framing devices for the act of diagnosis. Aligning the model with the universal model of diagnostic reasoning proffered by *Croskerry*, we can suggest that this meta-model is representative of the calibration stage of the diagnostic process. As a means to further decision support, the model provides a representation of the information entered into the electronic medical record through narrative (text). This representation is capable of providing feedback for diagnostic decision making and documentation of the practicing physician.

## Data Availability

The dataset used by this study was printed by the W. B. Saunders Company, now an imprint of Elsevier, the availability is the same as that of the printed book.

## Declarations

### Ethics Approval

This study did not require institutional review board approval, as it did not meet the requirements of human subjects research.

### Consent for publication

Not applicable.

### Author’s contributions

DC developed the initial theory, and performed the linguistic analysis. MBR, SMD, JB and MD contributed to the refinement of theory and validation of the linguistic analysis. All authors discussed, read and approved the manuscript.

## Acknowledgements

Not applicable.

## Appendix A: Appendix

### Figures

**Figure 23.**
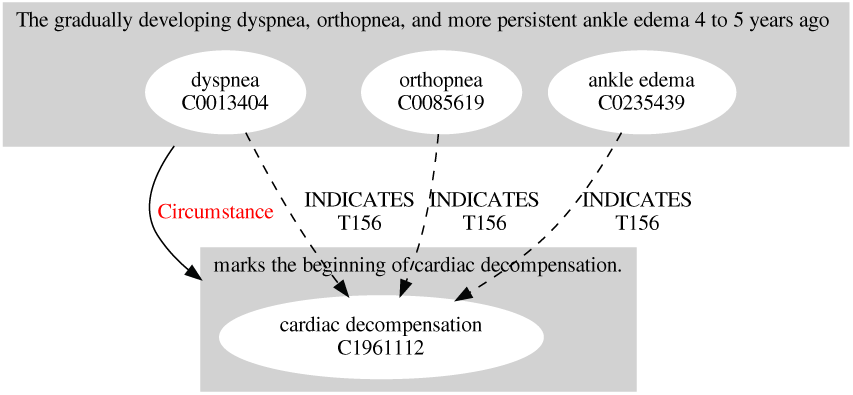
Propositional and Structural Semantic Perspectives Concept Integration. For topic sentence 1c, integrating Figures 11 and 17.

**Figure 24.**
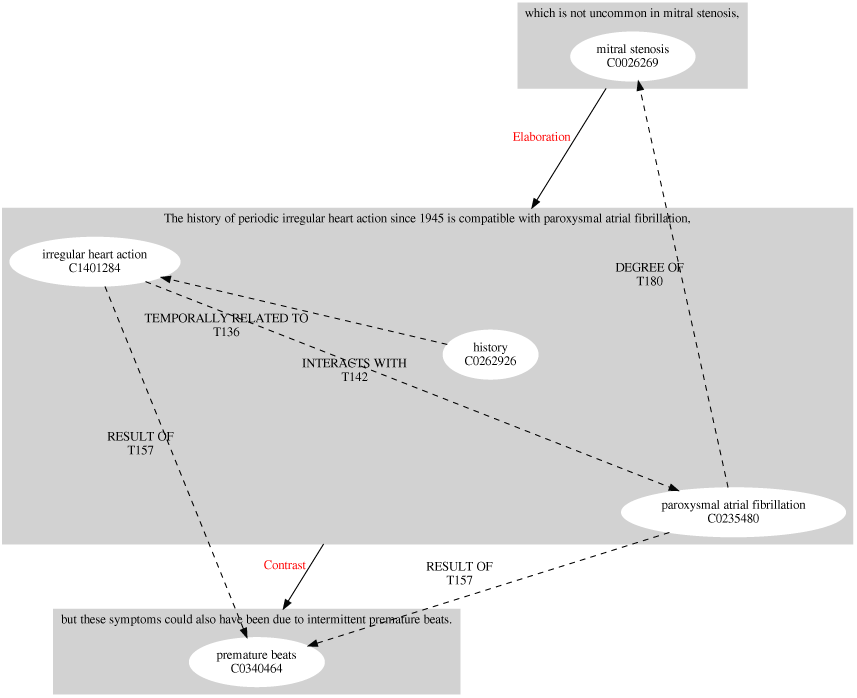
Propositional and Structural Semantic Perspectives Concept Integration. For topic sentence 2a, integrating Figures 12 and 18.

**Figure 25.**
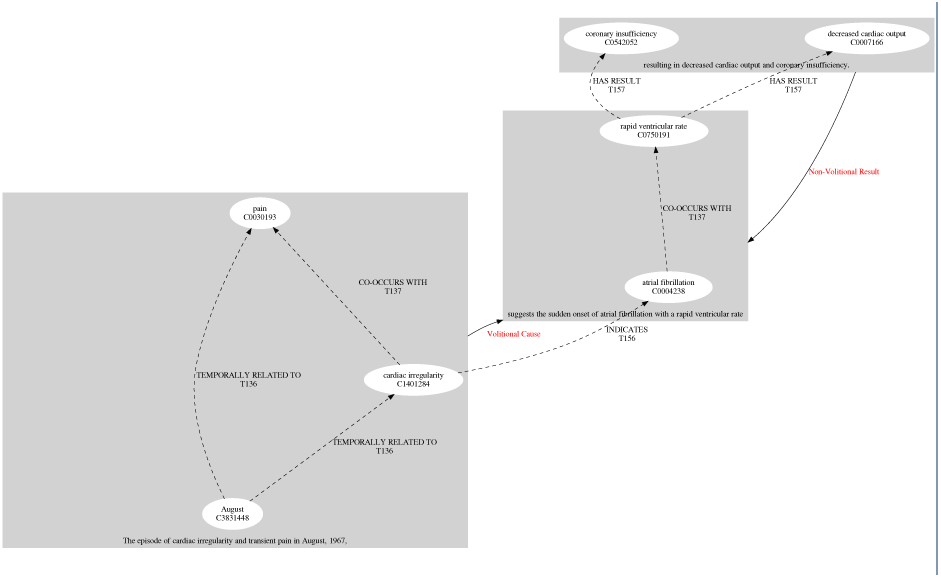
Propositional and Structural Semantic Perspectives Concept Integration. For topic sentence 2b, integrating Figures 13 and 19.

**Figure 26.**
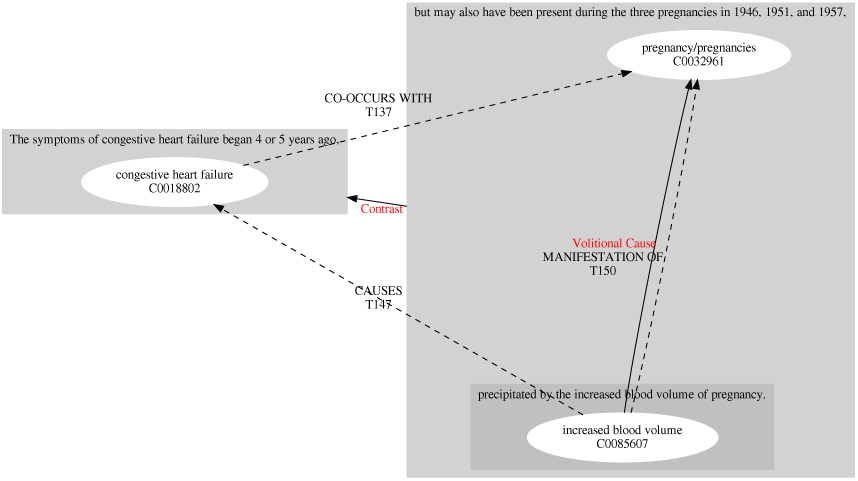
Propositional and structural semantic Perspectives concept Integration. For topic sentence 3, integrating Figures 14 and 20.

**Figure 27.**
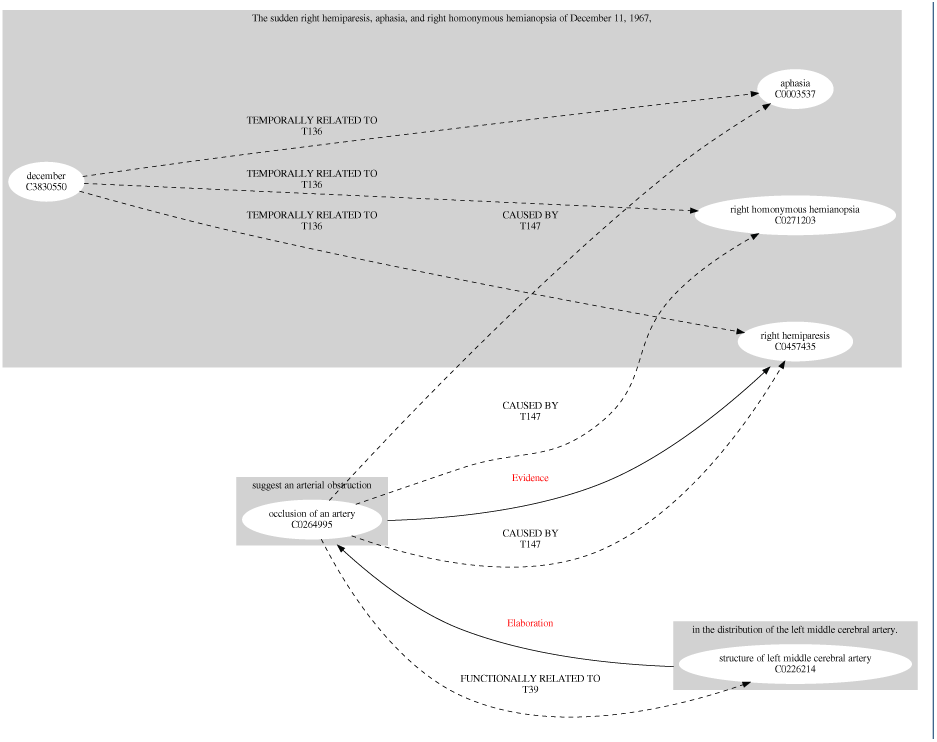
Propositional and structural semantic Perspectives concept Integration. For topic sentence 4, integrating Figures 15 and 21.

## Footnotes

[1] Retrieved from Trésor de la Langue Française informatisé, via the Analyse et Traitement Informatique de la Langue Française project. <http://stella.atilf.fr/>

